# From raw data to a score: Comparing quantitative methods that construct multi-level composite implementation strength scores of family planning programs in Malawi

**DOI:** 10.1101/2021.10.21.21265134

**Authors:** Anooj Pattnaik, Diwakar Mohan, Scott Zeger, Mercy Kanyuka, Fannie Kachale, Melissa A. Marx

## Abstract

**Background:** Data that capture implementation strength can be combined in multiple ways across content and health system levels to create a summary measure that can help us to explore and compare program implementation across facility catchment areas. Summary indices can make it easier for national policymakers to understand and address variation in strength of program implementation across jurisdictions. In this paper we describe development of an index that we used to describe the district-level strength of implementation of Malawi’s national family planning program.

**Methods:** To develop the index, we used data collected during a 2017 national, health facility- and community health worker Implementation Strength Assessment survey in Malawi to test different methods to combine indicators within and then across domains (4 methods – simple additive, weighted additive, principal components analysis, exploratory factor analysis) and combine scores across health facility and community health worker levels (2 methods – simple average and mixed effects model) to create a catchment area-level summary score for each health facility in Malawi. We explored how well each model captures variation and predicts couple-years protection and how feasible it is to conduct each type of analysis and the resulting interpretability.

**Results:** We found little difference in how the four methods combined indicator data at the individual and combined levels of the health system. However, there were major differences when combining scores across health system levels to obtain a score at the health facility catchment area level. The scores resulting from the mixed effects model were able to better discriminate differences between catchment area scores compared to the simple average method. The scores using the mixed effects combination method also demonstrated more of a dose-response relationship with couple-years protection.

**Conclusions:** The summary measure that was calculated from the mixed effects combination method captured the variation of strength of implementation of Malawi’s national family planning program at the health facility catchment area level. However, the best method for creating an index should be based on pros and cons listed, not least, analyst capacity and ease of interpretability of findings. Ultimately, the resulting summary measure can aid decisionmakers in understanding the combined effect of multiple aspects of programs being implemented in their health system and comparing strengths of programs across geographies.

## Background

Implementation strength assessments (ISAs) measure the intensity with which packages of interventions are delivered [1-3]. Results from ISAs indicate the amount of a program that is delivered, instead of how much of a program is received [4-6]. An ISA can give program managers and implementers specific information about what is and isn’t working in their program so they can make real-time improvements.

Quality of care (QoC) frameworks include implementation strength. In general, ISA fits into the Donabedian framework and its three dimensions of structure, process, and outcomes, and specifically for family planning, the Bruce-Jain framework divides QoC into six elements of FP programs [7-9]. Strength domains for this family planning (FP) assessment focus on the structural side of these frameworks, and include training, supervision, FP method choice and availability, demand generation activities, and accessibility [7].

In 2017, as part of the National Evaluation Program (NEP), the National Statistics Office (NSO) of Malawi conducted an ISA to understand the intensity of implementation of their national family planning program [10]. Studies similar to this type of evaluation, where the output of multiple programs rather than a single one is evaluated were reviewed to inform the design of this evaluation [5,6,11,12]. Yet, there has been limited ways to summarize the strength of large-scale, multi-pronged FP programs being implemented in low and middle-income countries into measures that can be analyzed against FP program outcomes and impacts.

In Malawi as in many low and middle-income countries (LIMC), the FP program includes both programs implemented at the facility and community levels. Malawi’s Ministry of Health, Christian Health Association of Malawi (CHAM), and NGO hospitals and health centers all provide healthcare services, including family planning. FP services are delivered by different types of health care workers: the health facility in-charge nurses, (“ICs”), health facility workers (HFWs), and two sets of community health workers: Health Surveillance Assistants (HSAs), and Community-based Distribution Agents (CBDAs) [13]. Community health workers (CHWs) are critical parts of the family planning and health system in Malawi. In particular, HSAs were able to provide the most popular method of family planning in the country as of 2017: injectables [14]. More broadly, the literature shows that CHWs are an essential source for FP methods and demand generation, especially in low-income settings [15-17].

When assessing the performance and strength of program implementation, it is important to consider how the data will be used. While detailed, granular results of these evaluations are valuable for implementers to use to improve their programs, national experts and policy makers are interested in understanding how strength of program implementation relates to impact and varies within and across countries. They may benefit from having a quantitative measure of strength of implementation that takes into account multiple domains and health system levels. The construction of summary measures can be a valuable way to facilitate more complex exploration [18-20].

Previous studies that summarize this type of data, often from Service Provision Assessments (SPA), have used four summary measure methods: simple additive, weighted additive, principal components analysis (PCA), and exploratory factor analysis (EFA) [21-25]. These studies have reported results at either the health facility or the community health worker (CHW) level, but not a combination of facility and CHW strength as a single index.

Drawing from studies in other fields, we found one simple and one more complex way to combine multiple levels of data: (i) aggregating lower-level data up to the higher level or, (ii) using a Bayesian mixed effects model (MEM). The benefit of the MEM is that it uses prior information to produce a posterior distribution of more accurate IS scores and can also account for clustering at multiple levels [26,27].

This study explores multiple ways implementation strength data can be combined across content and health system levels to create a summary measure for a facility catchment area (CA) and the advantages and disadvantages of each.

## Methods

### Data collection

Methods of the 2017 Malawi ISA, including the sampled population, their background characteristics, and findings for each IS indicator have been reported previously [7]. Briefly, in 2017 we measured the quantity of FP programs delivered in Malawi across the five IS domains (Table 1) by interviewing by mobile phone In-Charge Nurses (IC), health facility workers (HFW), health surveillance assistants (HSA) and community-based distribution assistants (CBDA) in all 28 districts. Interviews were conducted with workers associated with 660 (of all existing 666) health facilities including 602 ICs, 1662 HFWs, 4131 HSA, and 3187 CBDA.

**Table 1.** Indicators per implementation strength domain and health worker type

| IS Domain | Indicator | HW Type |
| --- | --- | --- |
| Training | Appropriately trained in FP* | HFW, HSA, CBDA |
|  | Ever trained in YFHS | HFW, HSA, CBDA |
| Supervision | Supervised for FP in last 3 months | HFW, HSA, CBDA |
|  | Last supervision covered youth FP topics | HSA, CBDA |
|  | HF has received supervision that included FP from someone external to the facility in previous 3 reporting months | IC |
|  | HFs whose supervision checklist of HWs includes Youth FP | IC |
| Contraceptive Methods and Supplies | Provides range of FP methods appropriate to type** | IC, HSA, CBDA |
|  | Appropriate FP method available on day of interview** | IC, HSA, CBDA |
|  | Has FP guidelines and job aids | IC, HSA, CBDA |
|  | Has youth FP guidelines | IC, HSA, CBDA |
|  | HF provides FP methods branded with social marketing | IC |
|  | HF has FP pamphlets | IC |
| Demand Generation Activities | Conducted youth event in last 3 months | IC, HSA, CBDA |
|  | Conducted SRH talks in last 3 months | HSA, CBDA |
|  | Conducted youth spaces in last 3 months | IC, HSA, CBDA |
|  | Conducted community meetings in last 3 months | IC, HSA, CBDA |
|  | HF has peer educators for FP | IC |
| Accessibility | Ensures privacy during FP consultations | IC, HSA, CBDA |
|  | Provides FP at least more than 12 hours per week | HSA, CBDA |
|  | Provides FP at least more than 24 hours per week | IC |
|  | HF has private room for FP consultations | IC |
|  | HF has space designated for youth consultations & activities | IC |
|  | HF has conducted mobile outreach since Jan 2017 | IC |

\*Pertains to whether the HW is appropriately trained out of the choices of counseling, condoms, OCPs, injectables, and implants. HFWs should be trained in all, HSAs on all except implants, and CBDAs on all except injectables and implants
\*\*Same as appropriate training. Provision and availability of method type is based on HW type.

All interviews were conducted from a call center in Zomba from April to August 2017 by trained interviewers. Interviews followed structured questionnaires and observation tools, and each type of health worker answered only questions relevant and appropriate for his or her type (Table 1). We summarized results and presented them to local decision-makers.

### Creating a summary score: Selection of Indicators

As depicted in Figure 1, we tested different methods to create a catchment area-level IS summary score for each health facility in Malawi. To do this we (i) combined indicators within and then across domains using 4 methods and (ii) combined scores across health system: facility and CHW, levels using 2 methods.

**Figure 1.**
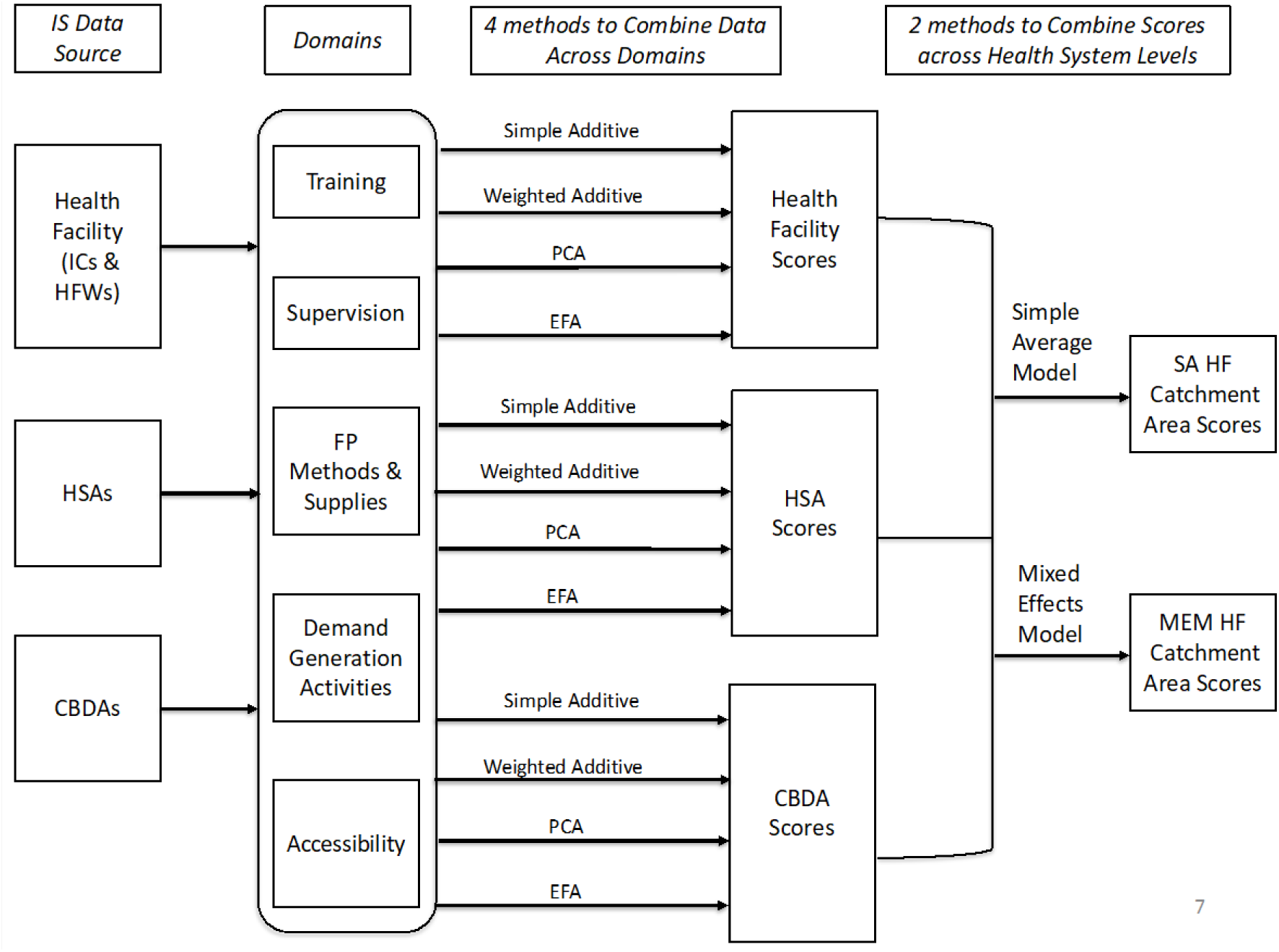
How scores are combined across domains and health system levels to construct implementation strength summary measures

### Combining indicators within and across domains (4 methods)

#### Simple Additive Summary measure (SA)

For the simple additive method, we first used an a priori hypotheses to narrow to the key sentinel indicators based on theory and expert input. Then we added all IS indicators to obtain a total score with equal weighting for each selected indicator. The total score was then divided by the total number of indicators to give a SA score.

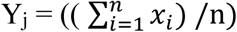

Where x is the indicator and n is the total number of indicators.

#### Weighted Additive Summary measure (WA)

Similar to SA, for the weighted additive method, each indicator within a domain is added together and then divided by the sum of indicators in that domain. These domain scores are added together and then divided by the total number of domains, creating the total weighted additive score.

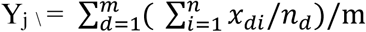

Where d refers to domains and m is the total number of domains.

#### Principal Component Analysis (PCA)

The PCA is a way to combine across the ISA indicators by reducing the highly correlated indicators into a smaller set of uncorrelated principal components that maximizes the amount of variation in the data. These components serve as analogues to the domains in the additive models above. The PCA uses all the IS indicators rather than being more parsimonious of choosing indicators as in the additive indices. We determined the number of components to use via parallel analysis, which is the most common method for doing this [28]. We then selected the indicators with loadings above 0.3 per convention and used them as weights for each indicator [29].

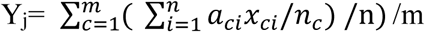

Where i is the number of indicators; a represents the factor loadings of each indicator for each jth health worker or facility; c is components; m is total number of components; y is equal to the predicted score from the chosen components for each jth health worker or facility

#### Exploratory Factor Analysis (EFA)

EFA is a variable reduction technique similar to a PCA, but it hypothesizes an underlying relationship among the set of IS variables. In this way, it estimates factors (as opposed to components) that account for only the common variance in the data, as opposed to the components in the PCA which reduces the dimensions of the data using total variance of the observed variables [30].

The components and factors emerge from the statistical analysis in PCA and EFA, whereas the domains are constructed a priori in the additive indices. As with the PCA, for EFA the full set of indicators is included, a parallel analysis determines the number of factors to use, and the factor loadings above 0.3 are kept and used as indicator weights. The equation is the same as the PCA, with the new factor loadings replacing the component weights. Cronbach’s alpha for internal consistency was calculated for the individual summary measure items.

Each of these four methods combine data across indicators separately at the health facility and CHW levels, resulting in two sets of scores for each method. The next sub-section describes how these scores are then combined across the facility and CHW levels; the last phase depicted in Figure 1.

#### Combining sxores across health system levels (2 methods)

We used two established methods to combine scores that come from multiple levels of data: the strength contributed by the facility itself (IC), by health facility workers (HFW) and by community-level workers (HSA and CBDA) into an overall measure of strength for the catchment area. In our case, the scores at the health facility and health worker levels (created from the four methods above) are used to model IS at the catchment area level. The first method was a simple, averaging model and the second was a more complex, Bayesian mixed effects model. They provide the option of a simple and a more complex method, from which analysts can choose.

A total of 8 different indices are compared: 4 indices (SA, WA, PCA, EFA) where facility and worker data are combined using the simple average method, and 4 indices (SA, WA, PCA, EFA) where we use the mixed effect model.

#### Simple average model

For the simple option, we constructed the IS score at the catchment area level by using three steps. First, we calculated an IS score at the HSA/CBDA level using the four methods described above. Then, we calculated the IS score at the health facility level by combining indicators from the IC and HFW surveys. If a health facility had multiple HSAs or CBDAs, the HSA and CBDA scores were averaged separately up to the facility level and added to the facility score as two extra (HSA average + CBDA average) domains in this model. For the scores using the simple and weighted additive methods, 2 domain scores (one for HSAs and one for CBDAs) were added to 14 indicators across 5 domains from the facility level. These scores are treated simply as indicators and added to the other 14 for the simple additive method. For the weighted average method, we treated these scores as domains and added them to the other 5 domain scores. The same process was used for the PCA and EFA indices, moving from CHW to facility to catchment area scores. The aggregated HSA/CBDA indicators with factor loadings above the threshold are included in the PCA or EFA.

#### Bayesian mixed effects model

We also used a Bayesian mixed effect model by using the scores for each individual health worker and health facility as the prior distribution to produce a posterior distribution of IS scores at the catchment area level. This approach benefits from the ability to borrow information from similar health facilities and workers to construct a representation of IS across a facility’s catchment area.

We created a three-level random effects model with individual health workers nested within facilities, which were nested within districts. The fixed effects were health facility type (hospital or health center), managing authority of the facility (MoH, CHAM, NGO), region (North, Central, South), and a dummy variable called “level” that designated whether the data was for an individual health worker or health facility. The outcome was the IS score from one of the four summary measure options. Different model specifications (for fixed and random effects) were compared with respect to model fit (Akaike Information Criteria, AIC, and log likelihood) and the percentage of variance explained [30,31].

#### Comparing summary measures

The resulting eight score distributions were compared using two-way scatterplots, box plots, kappa statistic scores, and funneling plots. To better understand the criterion validity of each summary measure, we also modeled couple-years protection (CYP) for each measure. CYP estimates the amount of protection provided by FP services over the course of a one-year period based on the volume and type of modern contraceptives provided and is calculated by multiplying the quantity of each modern method reported to have been used by a conversion factor. Each contraceptive method type has a different conversion factor (e.g., condoms are 120 units per CYP; injectables are 4 doses per CYP). The calculations yield estimates of the duration of protection provided by one unit of each contraceptive method which are added together to obtain a total CYP [32,33]. CYP was calculated using service utilization data collected in the 2017 Malawi ISA from health facilities and CHWs.

We divided the catchment area IS score distributions for each method into quintiles and analyzed how CYP changed as catchment areas scores in each IS quintile increased. Because we measured IS at the catchment area level, we adjusted CYP by the population of each catchment area. Otherwise, catchment areas with larger populations could have larger CYPs that could skew the relationship between IS and CYP at the catchment area level. The data source for catchment area population was the 2008 Malawi population census report [34].

In this way, we aimed to compare methods on how to combine data across content domains and across health system levels with the ultimate aim of creating a score for the implementation strength for the entire catchment area of each health facility in Malawi. We tested not only how well each model captures variation but also factored in how technically complex it is to conduct each type of analysis and whether different key audiences could easily interpret results.

All the analyses were conducted using R version 3.4.1 software [35]. The Johns Hopkins School of Public Health Institutional Review Board and the Malawi National Health Science Research Committee approved this study to collect the data in April 2017.

## Results

### Combining data across IS indicators and domains

Four methods were used to combine data across IS indicators per HW type, resulting in four sets of summary scores per HW type. Because questions asked of the in-charge nurse (IC) focused on structural quality/readiness of the facility itself while the questions answered by health workers (HFW, HSA, CBDA) focused on worker readiness, we combined the data considering the IC’s responses to represent the facility itself. Tables describing the factor loadings for each PCA and EFA model can be found in the supplementary section. Table 2 shows the median IS score and inter-quartile range (IQR) of implementation strength scores for each HW type and combination method. The median IS scores for the Health Facility (from the IC interview) range from 0.52 to 0.58 across all four combination methods, while HFWs range from 0.40 to 0.50. At the CHW level, IS scores for HSAs ranged from 0.45 to 0.49, while they ranged from 0.60 to 0.64 for CBDAs.

**Table 2.** Median and interquartile range of implementation strength scores for each HW type across four methods to combine data across indicators

|  | Simple Additive |  | Weighted Additive |  | PCA |  | EFA |  |
| --- | --- | --- | --- | --- | --- | --- | --- | --- |
|  | Median | IQR | Median | IQR | Median | IQR | Median | IQR |
| Health Facility (IC) | 0.52 | 0.38-0.68 | 0.52 | 0.38-0.67 | 0.56 | 0.41-0.68 | 0.58 | 0.42-0.70 |
| HFW | 0.45 | 0.27-0.55 | 0.40 | 0.27-0.53 | 0.50 | 0.36-0.65 | 0.40 | 0.25-0.52 |
| HSA | 0.45 | 0.36-0.63 | 0.47 | 0.30-0.63 | 0.46 | 0.32-0.58 | 0.49 | 0.35-0.66 |
| CBDA | 0.64 | 0.45-0.73 | 0.60 | 0.43-0.73 | 0.61 | 0.47-0.73 | 0.64 | 0.49-0.76 |

### Combining health facility and CHW IS scores (2 methods)

#### Simple Average Combination Method

Figure 2 shows pairwise comparisons via two-way scatterplots of the distributions of each of the four sets of scores (SA, WA, PCA, EFA) that use the simple average method to combine across health system levels. All six of the comparisons resulted in a high correlation coefficient of above 0.93, showing that the distributions are similar with negligible variation.

**Figure 2.**
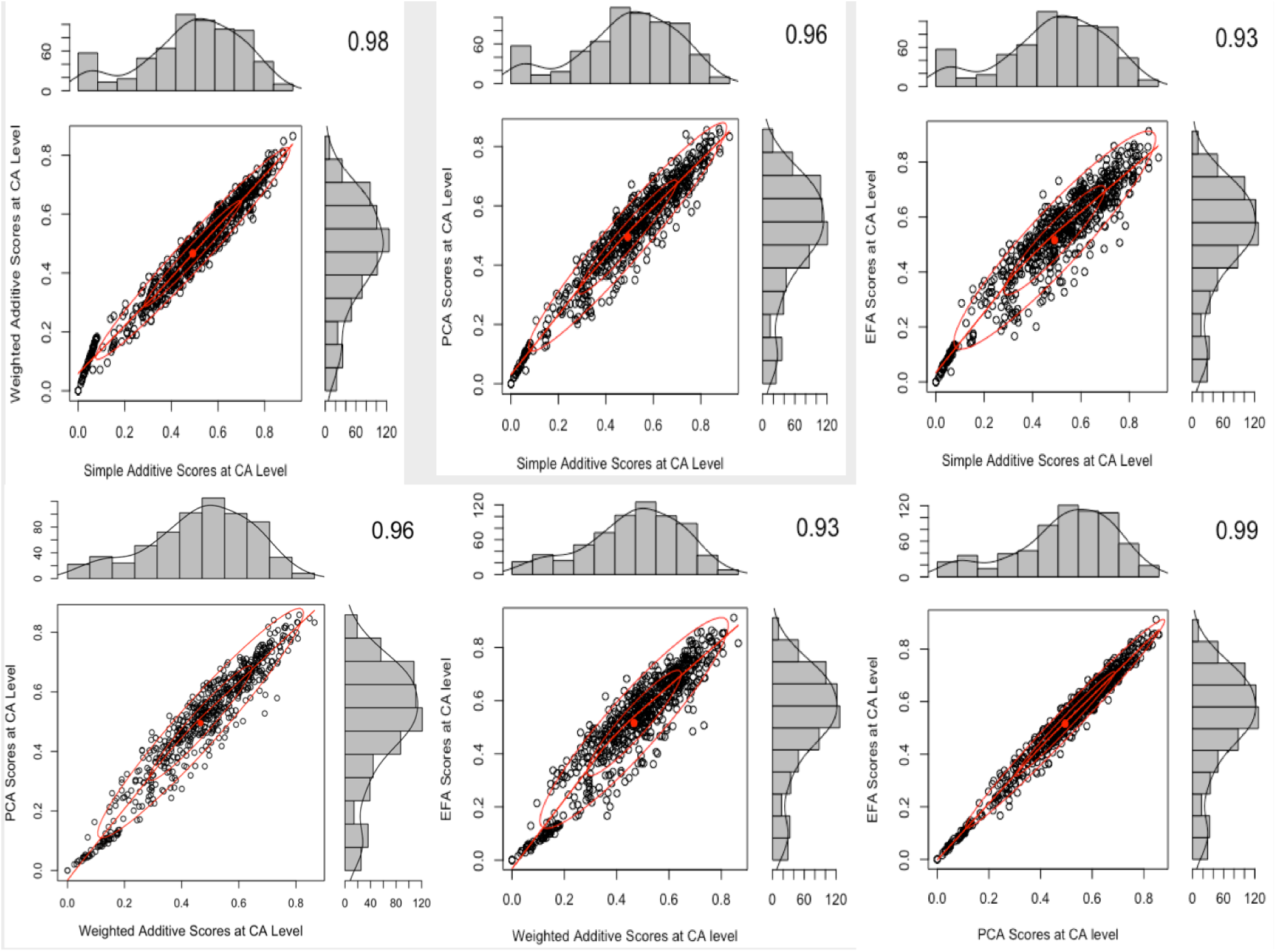
Two-way scatterplots comparing the four IS score distributions that use the simple average combination method

#### Mixed Effects Combination Method

We compared the fit of different regression models that included the IS score, facility type, managing authority of the facility, and the level dummy variable and chose the model with the best parameters. When the managing authority of the health facility was added as a fixed effect, variation at the facility level greatly reduced across the models. Results indicated that much of the variation at the facility level is confounded by whether the facility is managed by the Ministry of Health, CHAM, or an NGO. The models that used simple additive or exploratory factor analysis scores had the lowest model fit out of the four methods. The best model fit was the PCA model with the fixed effect of managing authority and the individual/facility dummy variable.

Figure 3 shows pairwise comparisons via two-way scatterplots of the distributions of each of the four sets of scores (SA, WA, PCA, EFA) that use the mixed effects method to combine across health system levels. Similar to the simple average score comparisons, there were high correlation coefficients across all six comparisons with little variation.

**Figure 3.**
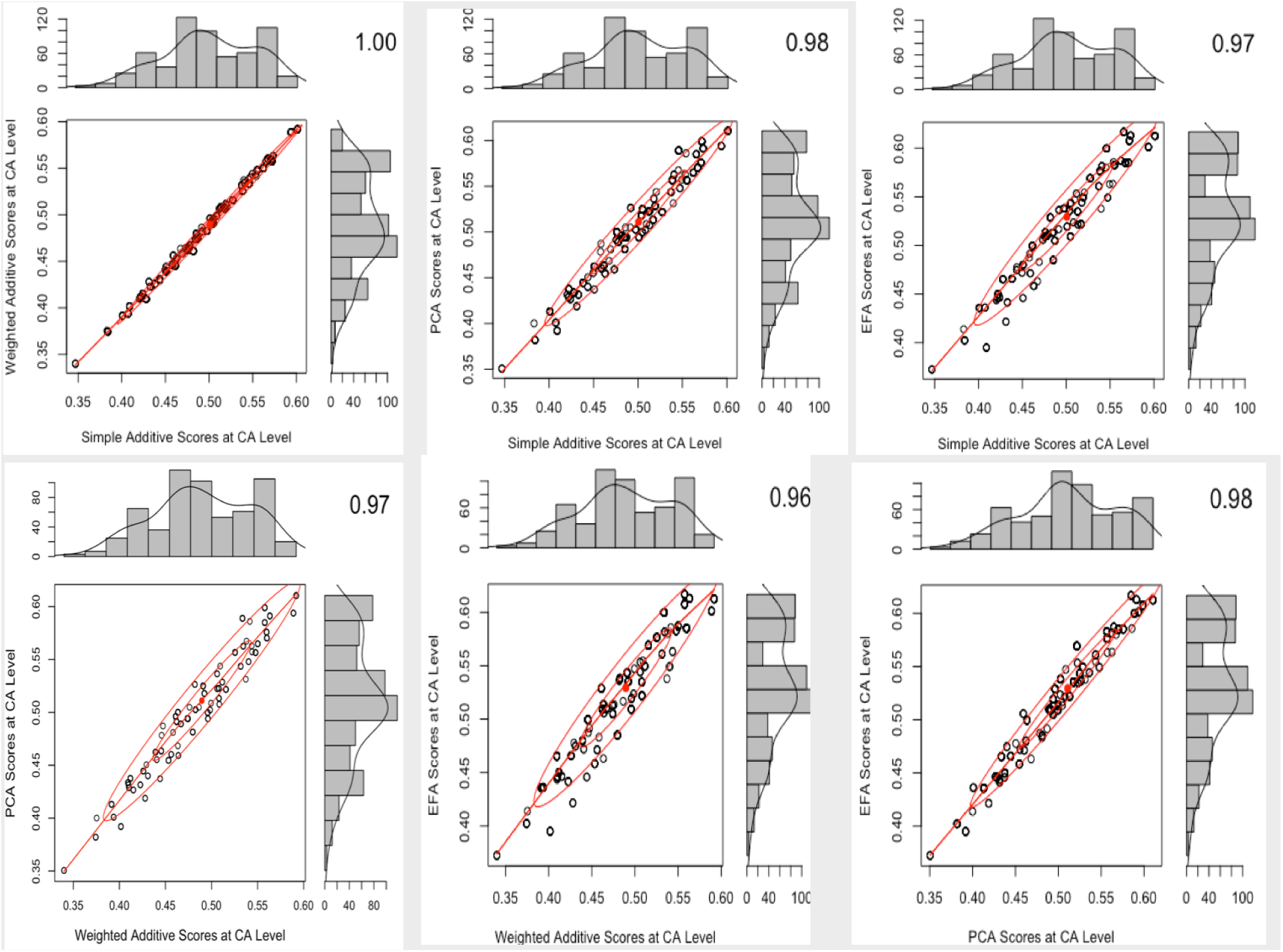
Two-way scatterplots comparing the four IS score distributions that use mixed effects combination method

#### Comparing score distributions between simple average and mixed effects methods

Figure 4 depicts a comparison of the CA scores that result from the simple average model and the mixed effects model, across the three regions in Malawi.

**Figure 4.**
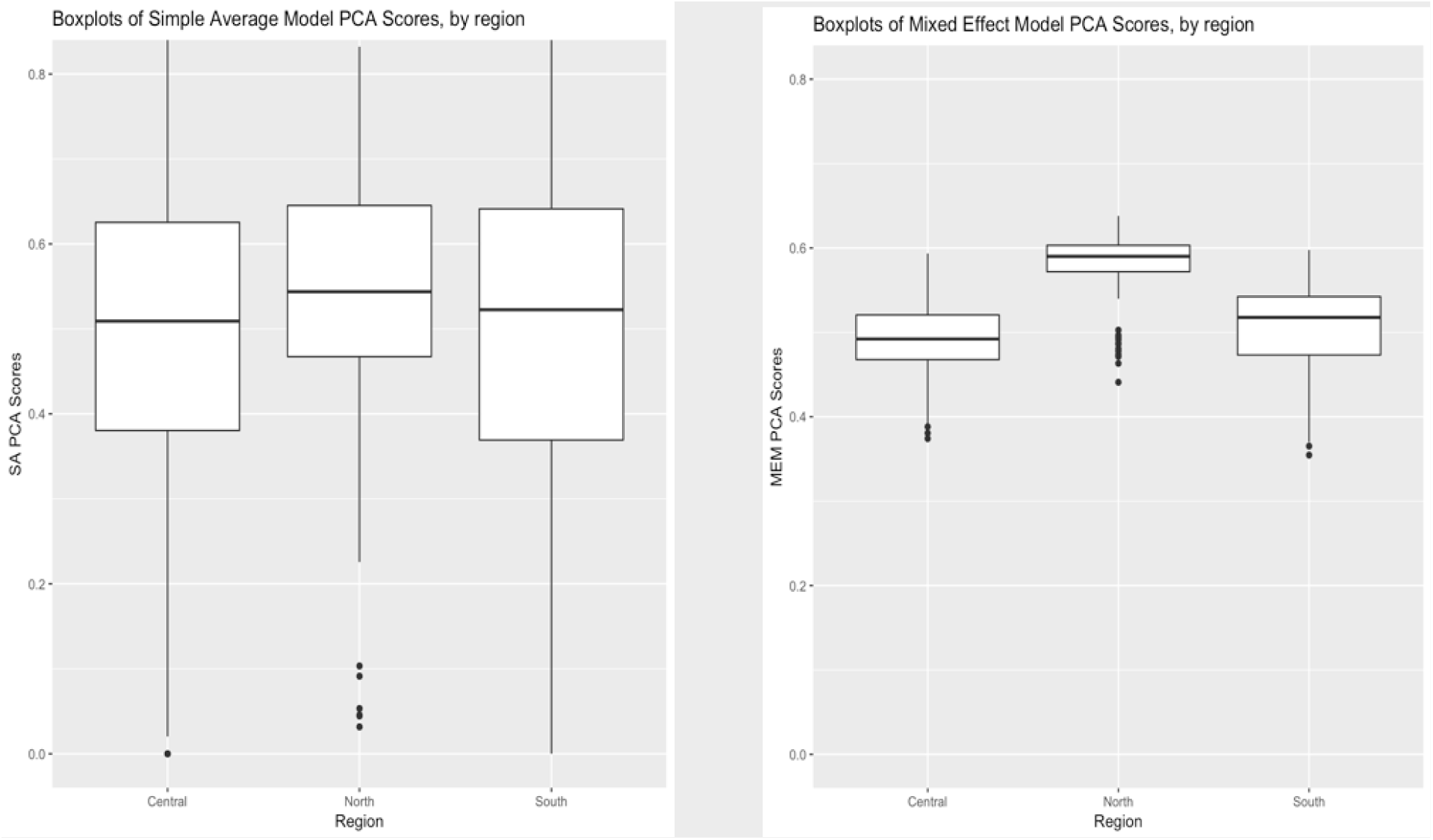
Comparison of mean and interquartile range of the ISA scores using the PCA summary method between the simple average and mixed effects model, by region

On the left, the range of scores resulting from the PCA using the simple average model are much larger than those resulting from the mixed effects model. In the diagram on the right, the uncertainty decreases dramatically, shown by the shorter boxes – and the difference between the regions are more obvious (and in two cases statistically significant). Essentially, MEM shrinks the distribution of scores and creates a more discriminatory set of scores than the simple average method.

Next, we compared the scores by dividing each score distribution into quintiles and comparing how each summary measure method ranks the facility catchment areas using a weighted kappa coefficient. For instance, was a catchment area that was ranked as a 2 using the weighted additive method and the simple average combination method, also ranked as a 2 when using the PCA method and the mixed effects combination model? Table 3 below shows the weighted Kappa coefficients between the simple average and mixed effects combination models. To interpret the strength of agreement for the kappa coefficient, Landis and Koch proposed the following standards: ≤0=poor, 0.01–0.20=slight, 0.21–0.40=fair, 0.41–0.60=moderate, 0.61– 0.80=substantial, and 0.81–1=almost perfect [36].

**Table 3.** Comparison of scores in quintiles between simple average and mixed effects combination models using weighted kappa

|  |  | *Simple Average Scores* |  |  |  |
| --- | --- | --- | --- | --- | --- |
| *MEM Scores* |  | SA | WA | PCA | EFA |
|  | SA | 0.31 | 0.31 | 0.26 | 0.27 |
|  | WA | 0.34 | 0.34 | 0.29 | 0.30 |
|  | PCA | 0.32 | 0.32 | 0.29 | 0.30 |
|  | EFA | 0.31 | 0.31 | 0.28 | 0.29 |

The level of agreement between the indices in Table 3 is only “fair.” On the other hand, weighted kappa coefficients comparing scores *within* each combination model were all in the “substantial” to “almost perfect” range. Similarly, the scatterplots shown earlier demonstrated strong correlations between the score distributions within each combination model. This finding lends further evidence that the four summary measure methods combining data across domains capture variation very similarly whereas the two methods that combine data across health system levels capture variation very differently.

Next, we assessed the criterion validity of each health system combination method by observing how the outcome variable of population-adjusted couple-years protection changes with increasing quintiles of implementation strength.

Table 4 depicts a dose-response relationship between IS scores and population-adjusted CYP in the mixed effects model scores, but no clear trend in the simple average combination model scores.

**Table 4.** Change in population-adjusted couple-years protection by quintiles of implementation strength, comparing simple average and mixed effects models

| Average<br>Population-Adjusted CYP |
| --- |

| IS Quintile | SA | MEM |
| --- | --- | --- |
| 1 | 16.97 | 27.17 |
| 2 | 49.42 | 30.16 |
| 3 | 38.33 | 30.47 |
| 4 | 36.16 | 36.73 |
| 5 | 49.24 | 65.52 |

## Discussion

Our findings indicate little difference in how the simple and weighted additive, PCA, and EFA captured variation of the IS data at the individual and combined levels of the health system. In fact, there was much higher agreement between the four methods in this study than the previous studies we reviewed that compared similar summary measures [25,37]. However, there were major differences when combining scores across health system levels to obtain a score at the health facility catchment area level, as reflected by the low weighted kappa coefficients comparing how each summary measure ranks facilities. The mixed effects model using Bayesian methods shrunk the variation catchment area IS scores. The scores resulting from the mixed effects model better discriminated differences across the regions compared to the simple average method. The scores using the mixed effects combination method also demonstrated more of a dose-response relationship with CYP than the simple average method.

There are several factors to consider when choosing between the four methods that combine across IS domains and indicators. While the simple and weighted additive methods are relatively easy to calculate and interpret, they have a number of limitations. The additive measures heavily rely on a priori input from experts in choosing what domains and indicators should be included and how they should be grouped. Future studies should consider a rigorous process of expert input, such as a Delphi method, to decide which indicators are included [22,38]. The simple additive method assigns equal weights to each indicator and could over- or under-weigh certain indicators. It also does not account for collinearity among indicators or across domains [22]. The heaping of scores in the distributions present challenges in the utility of the simple additive method [23,44]. The weighted additive method can address some of these concerns by accounting for collinearity within a domain.

Using a PCA or EFA to combine data across IS indicators is more complex to calculate and more difficult to interpret than the additive options. There are several considerations (e.g., factor extraction, rotation, components to retain) in constructing the score from a PCA or EFA that require a strong understanding of the method [39,40,42]. Yet, the weighting challenges in the additive methods are not applicable to these factor analyses. The number of components or factors to retain, which serve as analogues to the domains in the additive models, come from the underlying variation of the data itself. A drawback of PCA and EFA scores is that they are empirical [22,45,46]. The weights for the indicators derived from the PCA and EFA are only applicable the dataset under consideration. Another drawback for the EFA is that it requires a priori decisions about the composition of the domains and indicators [41,42,46].

There were few relevant published studies in the context of global health and implementation science to guide our choice of combining data across different health system levels. Several factors should be considered when deciding between the simple average and mixed effects methods to combine data across health system levels. As the name suggests, the simple average technique is easier to construct. However, this method can lead to potentially less precise catchment area scores, especially when there are fewer CHWs per facility. The mixed effects model (MEM), on the other hand, uses prior information from similar health facilities and workers to reduce the influence of extreme values. The MEM scores were better able to discriminate regional differences and had a clear dose-response relationship with CYP than the simple average scores. Mixed effects combination method consistently outperformed the simple average method in our study.

This study explored different ways to create a composite score for how strongly multiple large-scale family planning programs are being implemented across different health system levels. Many studies reviewed used the four methods (simple and weighted additive, PCA, EFA) to combine data across content areas and used multi-level modeling in FP or maternal and child health research, but none that we encountered combined facility and community level data to create a summary level measure at the catchment area level [43-45]. Several studies took the health system into account by including a single indicator for whether a facility had CHWs in the construction of their facility-level summary measure [45,46]. Our study accounts for CHW contribution more comprehensively and explicitly by creating separate scores for individual CHWs and then combines them with the facility. Many other studies analyze individual indicators or construct summary measures for each individual domain, rather than one across multiple domains [46-48]. For instance, other studies may want to explore the effect of a specific intervention that trained HSAs in Malawi on YFHS. Our study explored options to combine data both across indicators/domains and health system levels.

Summary measures for IS can be used to understand the combined impact of a set of FP programs, identify variations in implementation across geographic areas, and assist with targeting priority areas for future implementation. For instance, district leadership can review the IS scores across facilities in the geographies in their jurisdictions to quickly identify where performance differs to identify the cause and prioritize additional resources and/or interventions in response. However, the score does not indicate why strength is low or high and must be supported by a deeper dig into the data and/or possibly additional data collection. Repeated application of the ISA can also allow policy experts and program implementers to track the trend in IS over time. Due to the relatively simplicity of the types of questions that can be asked via short phone interviews or using routine data, tracking IS scores across time can give decision-makers a valuable tool in rapidly assessing progress towards their objectives.

These summary measures could also be compared across countries to better understand how the strength of implementation of FP programs varies from one context to another. An analogue to this is the Family Planning Effort Index, where 10-15 key informant respondents in each country respond to a questionnaire that gauges the country’s FP effort levels, and results in a score [19]. The ISA score resulting from the methods explored in this study come from combining a much larger quantity of input and process level data across a wider range of domains and health system levels, not just stakeholder input. Future studies could produce these ISA scores in countries other than Malawi and explore how these IS scores would change in different contexts and systems. In turn, national policymakers and international experts can use these scores to better understand how strongly national FP programs are being implemented and the relative strength of implementation between different countries.

There is a need to explore how the construction of the scores would change if applied in other areas, such as maternal and child health programs. Future research can also explore the associations between the IS scores and key FP outcomes further down the impact chain than CYP, such as modern contraceptive prevalence rate (mCPR) and demand satisfied for FP. The ultimate and explicit objective of these FP programs is to positively impact these outcomes down the impact chain [4].

### Limitations

The data and indicators used were limited to those from the 2017 Malawi ISA. These indicators may not capture every possible indicator related to the implementation of every FP program in Malawi. Still, the study aimed to capture the major interventions after review of the local policies and input from local leadership in the Ministry of Health, CHAM, and leading NGOs. The ISA does not capture quality of care received; for instance, even if a health facility has a high IS score, its health workers could be providing poor quality care in person to the client. Still, capturing structural quality is important and the indicators are often more easily measurable [1,2,7,10]. There could also be more ways to combine information across content and health system levels that were not explored in this study. After a careful review of the literature, we aimed to choose the most common methods used, as well as a range of methods from simple to more complex.

We adjusted CYP (calculated from the 2017 Malawi ISA) by catchment population data from the 2008 Malawi census, which was collected nearly a decade before the ISA. Population-adjusted CYP as calculated in this study is likely overestimated due to the likely increase in the population in each catchment area over the decade. The CYP should be readjusted using the 2018 Malawi census report, which had not been released at the time of writing this paper. Also, when deciding which method is best, we assume that CYP should be associated causally and in a dose-response relationship with strength of family planning programs. The literature is mixed on the role strength (aka intensity, structural quality) plays in family planning outcomes.

### Conclusions

This study lays out a roadmap on how to construct a summary measure for implementation strength of large-scale programs, combining across different levels of a health system. It can serve as a guide for researchers aiming to construct their own composite scores or indices, because it clarifies the pros and cons of each method choice and provides options based on technical capacity. It can also aid decisionmakers in understanding the total effect of multiple programs being implemented in their health systems. It can then serve as an evidence-based platform to target areas with weaker implementation, especially in low and middle-income contexts where resources and capacity may be constrained.

## Data Availability

All data produced in the present study are available upon reasonable request to the authors

## List of Abbreviations

AIC: Akaike information criteria
CA: Catchment area
CBDA: Community-based distribution agent
CHAM: Christian Health Association of Malawi
CHW: Community health worker
CYP: Couple-years protection
EFA: Exploratory factor analysis
FP: Family planning
HFW: Health facility worker
HSA: Health surveillance agent
IC: In-Charge (of a health facility)
IQR: Inter-quartile range
IS: Implementation strength
ISA: Implementation strength assessment
LMIC: Lower and middle income country
mCPR: Modern contraceptive prevalence rate
MOH: Ministry of Health
MEM: Mixed effects model
NEP: National Evaluation Program
NGO: Non-governmental organization
NSO: National Statistics Office
PCA: Principal components analysis
QoC: Quality of care
SA: Simple additive
SPA: Service provision assessment
WA: Weighted additive
YFHS: Youth-friendly health services

## Declarations

## Acknowledgements

The authors would also like to thank the following organizations and individuals for their valuable support of this work: the RHD Unit of the Malawi Ministry of Health; the Malawi National Statistics Office, including Jameson Ndawala, Sautso Wachepa, Sam Chipokosa, and Lewis Gombwa; and the Institute for International Programs (IIP) including Neff Walker, Agbessi Amouzou, Jamie Perrin, and Emily Carter.

## Ethics Declaration

This study was approved by the Johns Hopkins School of Public Health Institutional Review Board and the Malawi National Health Science Research Committee to collect this health facility and worker data in April 2017.

## Availability of data and materials

Please contact author for data requests

## Authors’ Contributions

MM, DM, AP conceived and designed the study, with key input from SZ, MK, FK. AP led data analysis and wrote the first draft. MM, DM, and SZ commented on drafts. All authors contributed to interpretation of results and commented on drafts prior to publication. All authors read and approved the final manuscript

## Consent for publication

Not applicable

